# Modeling highlights the challenge of maintaining HCV micro-elimination among people who inject drugs

**DOI:** 10.1101/2025.07.25.25332212

**Authors:** Eric Tatara, Louis Shekhtman, Nicholson Collier, Scott J. Cotler, Mary Ellen Mackesy-Amiti, Basmattee Boodram, Harel Dahari, Marian Major, Jonathan Ozik

## Abstract

**Background & Aims:** People who inject drugs (PWID) are at high risk for acquiring and transmitting hepatitis C virus (HCV). Direct-acting antiviral (DAA) therapy leads to high cure rates. However, the lack of protective immunity after cure and high rates of reinfection in PWID necessitates access to multiple DAA treatments per PWID to reach the World Health Organization goal of HCV elimination, defined as 90% HCV incidence reduction. A major public health concern is that once the elimination goal is met, treatment of PWID will be stopped or significantly limited by a reduction in resources and complacency.

**Methods:** We refined and extended our agent-based model to study the effects of varying levels of DAA treatment among HCV-infected PWID from Chicago, IL and the effects of stopping or reducing DAAs treatment after elimination is reached. The model uses individual temporal viral load profiles to determine transmission probabilities relative to the HCV RNA titers of receptive syringe-sharing PWID.

**Results:** Modeling predicts that insufficient annual DAA treatment (≤2.5%, 25 per 1000 PWID) risks increasing HCV incidence. Elimination can be achieved within 10 years with annual treatment of ≥7.5%. When DAA treatment is stopped, the rate of new chronic HCV infections rapidly increases, exceeding the elimination goal within 12 months and returns to pre-elimination levels within 5 years. Annual treatment of ≥0.5% would maintain the elimination goal. This equates to identifying and treating 160 infected people in a PWID population of 32,000 each year, which would be highly resource intensive.

**Conclusion:** The challenge to maintain the elimination goal once met, underscores the importance of augmenting DAA treatment with interventions strategies such as syringe service programs and vaccines.

## INTRODUCTION

Worldwide >50 million individuals are estimated to be chronically infected with hepatitis C virus (HCV), with ∼1.0 million new infections occurring each year [1]. In the US alone an estimated 4 million people are infected with the virus [2]. In 2023, there were an estimated 69,000 acute cases and 11,194 HCV-related deaths. From 2013 to 2022 the incidence rate of acute hepatitis C more than doubled (126% increase) and increased 16% from 2019 to 2020, although for the first time in over a decade the number of acute HCV cases remained stable since 2021 [3]. Nonetheless, the recent transition to synthetic opioids with faster onset and shorter duration, may lead to more frequent injections and sharing of IDU [4]. Drug therapy for HCV has improved significantly in the last decade. Direct-acting antivirals (DAAs) successfully clear most infections with 8-12 weeks of treatment [5]. This prompted the World Health Organization (WHO) to endorse the ambitious goal of reducing HCV incidence by 90% and HCV-related mortality by 65% compared to 2015 levels by 2030 as part of their global health sector strategy for ending viral hepatitis worldwide [6]. Effective action plans have been introduced in several countries through price negotiations, community engagement, and education programs [7–9], and there is evidence of success in reaching the elimination goal in Egypt [9]. However, many countries still lag behind the elimination goals. The United States is behind on elimination targets, partly due to funding, the absence of a national registry of HCV-positive people, and systems to track those treated [7]. Between 2016 and 2021 the number of people in low and middle income countries receiving DAA treatment for chronic HCV infection increased almost 10-fold from the 2015 baseline of 1.1 million to 9.4 million [1]. However, treatment levels remained low, only 21% of people infected with HCV were diagnosed and 62% of those diagnosed received treatment. The report estimated that treatment coverage needs to increase nearly sixfold in the next decade to reach the 2030 targets [1]. Deficits in screening and diagnosis and linkage to care, high costs, restricted access, and the ongoing opioid epidemic that continues to fuel the HCV epidemic [10] have limited the impact of treatment. Although clearance of the virus through DAA treatment can restore dysfunctional immune responses that develop during chronic infections [11, 12], this is incomplete and treated people remain susceptible to reinfection and can develop new chronic infections [13–15]. For most countries, new strategies will need to be implemented to achieve the elimination goal [16], highlighting the importance of other measures to prevent HCV transmission.

Persons who inject drugs (PWID) are at high risk for acquiring and transmitting HCV through the sharing of contaminated syringes and drug paraphernalia [3, 17]. This population represents the major source for new infections and therefore the main target for testing and treatment to reach the WHO elimination goal. HCV elimination among PWID is a costly investment and understanding the levels of treatment needed to reach the goal can provide a more efficient pathway to screening and treatment. In addition, once the elimination goal is reached, the challenge of sustaining low incidence rates remains.

The three main goals of the current study were 1) to assess the level of treatment needed to achieve the WHO elimination goal within 10 years in a US PWID population, 2) the level of treatment that would be needed to maintain the reduced level of incidence once the goal is met and 3) to assess the impact on HCV incidence if funding for screening and treatment is withdrawn once the goal is met. To perform these studies, we extended our agent-based model (called HepCEP) developed using field data collected from Chicago-area PWID to create an in-silico population [18–21], which accounts for the geographic distribution of PWID, injection behaviors, syringe sharing networks, and new HCV infections. We added HCV in-host kinetics for all infected individuals during various infection stages [22] and DAA treatment [23] in order to represent the probability of transmission (as a function of temporal HCV titer) during each syringe sharing event [22].

We applied the extended HepCEP to study the effects of varying levels of treatment in DAA programs among HCV infected PWID on the time taken to reach the elimination goal, the level of continued treatment needed to maintain the goal once attained and the effects of stopping screening and access to DAAs once the goal is achieved.

## METHODS

### Extended Hepatitis C Elimination in PWID (HepCEP) Model

#### Model Population

The HepCEP model simulates the PWID population in Chicago Illinois, USA, and the expansive surrounding suburbs that span counties in Illinois, Wisconsin and Indiana. In this model, we included drug use and syringe sharing behaviors, and associated infection dynamics [18, 19, 24]. (See Supplemental File, Tables S2-S3). In brief, this includes data from enrollees of a large syringe service program (SSP) (n=6,000, 2006-13)[25], the Chicago injection drug use data collection cycles of the National HIV Behavioral Surveillance (NHBS) survey from 2009 (n=545)[26] and 2012 (n=209)[27], and other local studies [21, 28]. The prevalence of HCV RNA in the population is 31.6%.

The network evolves over time, and during the course of the simulation some connections (ties) may be lost, while new ties form, resulting in an approximately constant network size as previously described [18].

#### HCV Infection Dynamics using In-Host Viral Kinetic Models

The extended HepCEP includes a dynamic HCV state transition model for each individual PWID agent that determines HCV infection status after an infection occurs (Fig. 1). Individual viral load profiles were incorporated to better reflect the temporal dynamics of viral load and HCV transmission probability unique to each PWID. In addition, PWID undergoing DAA treatment were assigned a viral load time series reflecting lowered viral loads during treatment and ultimately returned to a susceptible state as cured individuals (sustained virological response, SVR) or rebound (non-SVR) to a chronic infection after treatment completion (Fig. 1). The viral kinetic profiles during acute infection, re-infection and DAA treatment (Fig. 2A-H and Table S1) were generated by HCV dynamic mathematical models [29–31] to allow the calculation of the likelihood of transmission as a function of viral load (Fig. 2I) [22]. These are described in detail in the Supplementary Material File.

**Figure 1.**
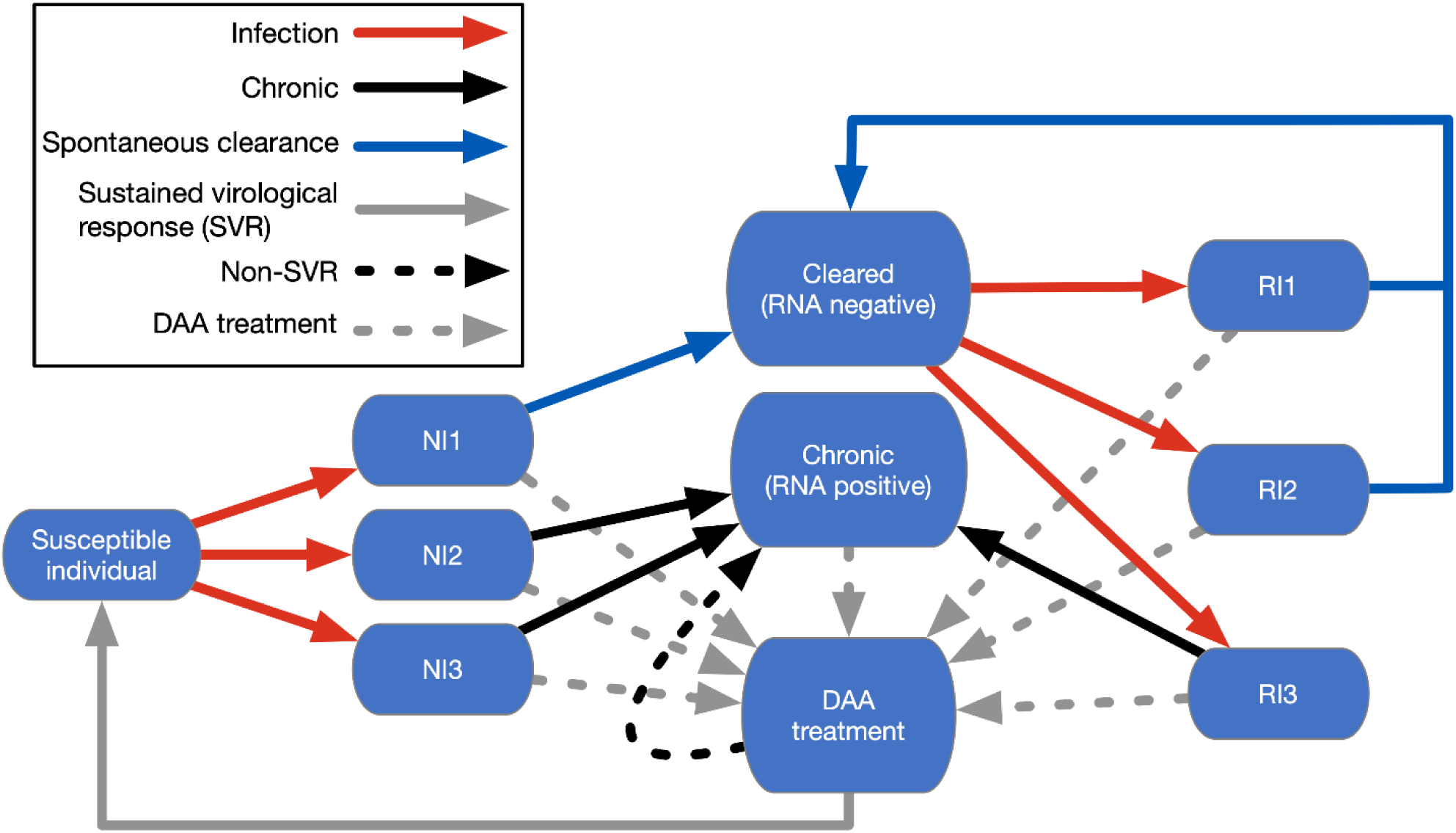
PWID agent HCV infection state transition model. Boxes represent HCV states, arrows represent transitions between states. Upon infection, naive individuals are placed into one of three infection states: acute with self-clearance (NI1) (Fig. 2A), acute with incomplete clearance (NI2) (Fig. 2B), acute persistent (NI3) (Fig. 2C). Reinfection results in agents transitioning to: reinfection with low titer (RI1) clearance (Fig. 2D), reinfection with high titer (RI2) clearance (Fig. 2E), chronic (RI3) (Fig. 2F), rebound long term (Fig. 2G) or DAA treated (Fig. 2H). Dashed gray lines: DAA treatment; Solid gray line: SVR; Black dashed line: non-SVR.

**Figure 2.**
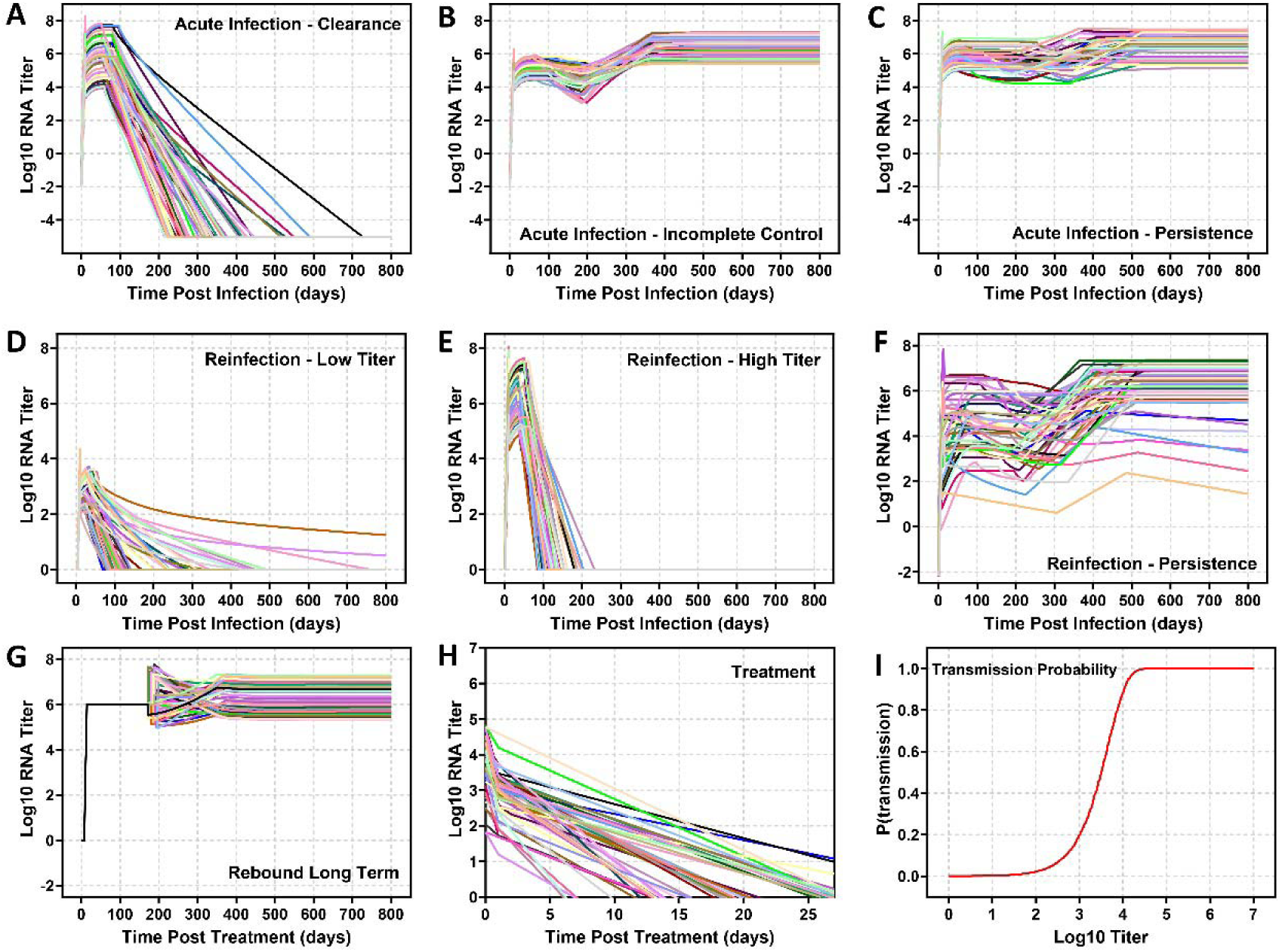
Patient in-host viral kinetic (VK) profiles and infection probabilities. **Acute infection (A) to (C):** Fifty different patient VK profiles for each acute infection group with **(A)** clearance, **(B)** incomplete control, and (C) persistence. **Reinfection (D) to (F):** Fifty VK profiles for reinfection with (D) low titer clearance, (E) high titer clearance, and (F) chronic reinfection. **Chronic infection (G):** Fifty possible long term VK rebound profiles. **Treatment profiles (H):** Fifty possible VK profiles during DAA treatment. (I) The likelihood of HCV transmission of needle/syringe-sharing donors as a function of viral load.

#### DAA Treatment

DAA Treatment was modelled as an unbiased random sampling of acute and chronically infected PWID at the annual target treatment rate, defined as the total number treated annually as a fraction of the total population, e.g. a 10% target rate (for 32,000 PWID) results in up to 3,200 PWID treated annually. The annual DAA target treatment rate is translated into a daily treatment quota such that the number of PWID treated annually is evenly distributed over the calendar year. The total PWID target for a single day was determined by the daily mean treatment level, which is the total PWID population multiplied by the annual treatment parameter / 365. The daily treatment target was sampled from a Poisson distribution using the daily mean treatment rate. This treatment process assumes perfect knowledge of the HCV state of the entire PWID population during a simulation and does not include efforts to screen PWID for HCV. Treatment duration was 12 weeks, the treatment adherence was fixed at 90% and the DAA treatment success probability was a function of the treatment adherence and SVR parameters. While recently reported SVR rates are close to 99% [32–34] in many populations, we used a conservative estimate for SVR rates of 90%. We assumed that successful treatment did not affect the probability of subsequent re-infections. Retreatment was allowed for PWID who successfully completed treatment and became re-infected.

#### Discontinuation and Reduction of DAA Treatment

Discontinuation of access to DAA treatment after elimination was modeled by changing the target treatment parameter to zero after the initial 10-year period for all target rates. To model reduced DAA treatment after elimination is achieved, we used a fixed 7.5% DAA target treatment rate for a 10-year period to first achieve elimination after which the DAA target treatment parameter was changed to a value from 0-7.5% for the remaining 20-year simulation time period.

### Agent-based Model Execution

The agent-based model (ABM) is run for 10 years to achieve steady-state baseline values for all model parameters after which a DAA treatment period was modeled for an additional 10-year period. The model was subsequently run for an additional 20-year period under different scenarios for sustained, discontinued, and reduced DAA target treatment rates.

We report the mean annual incidence of chronic HCV infections relative to the mean baseline incidence rate in year zero with no treatment (target rate of 0%). Each individual PWID agent steps through their current activities on each simulation day. Transitions between activities and HCV state is dependent on the agent’s current state (e.g., infected), associated viral load profile (if infected), and the types of activities, which include syringe-sharing events, syringe-sharing network connection formation and dissolution, and DAA treatment enrollment, DAA cure, and re-infection events.

Simulations were conducted using a high-performance computing workflow implemented with the EMEWS framework [36]. The simulation experiments were executed on the Bebop cluster run by the Laboratory Computing Resource Center at Argonne National Laboratory. Each simulation required approximately one hour of wall time to complete. Using the EMEWS workflow on the Bebop cluster, the actual compute time was also one hour since all runs could execute in parallel on 1,600 processes.

## RESULTS

### Insufficient DAA treatment reduces HCV RNA prevalence but increases incidence

When DAA treatment rates are as low as 2.5%, the relative incidence of new chronic infections rapidly increases above the pre-treatment baseline during the first years of treatment followed by a sustained increase at elevated levels 3-fold higher than the baseline for 30 years (Fig. 3A). In contrast, HCV RNA prevalence declines after DAA treatment starts with prevalence reaching ∼4% after 30 years (Figure 3B).

**Figure 3.**
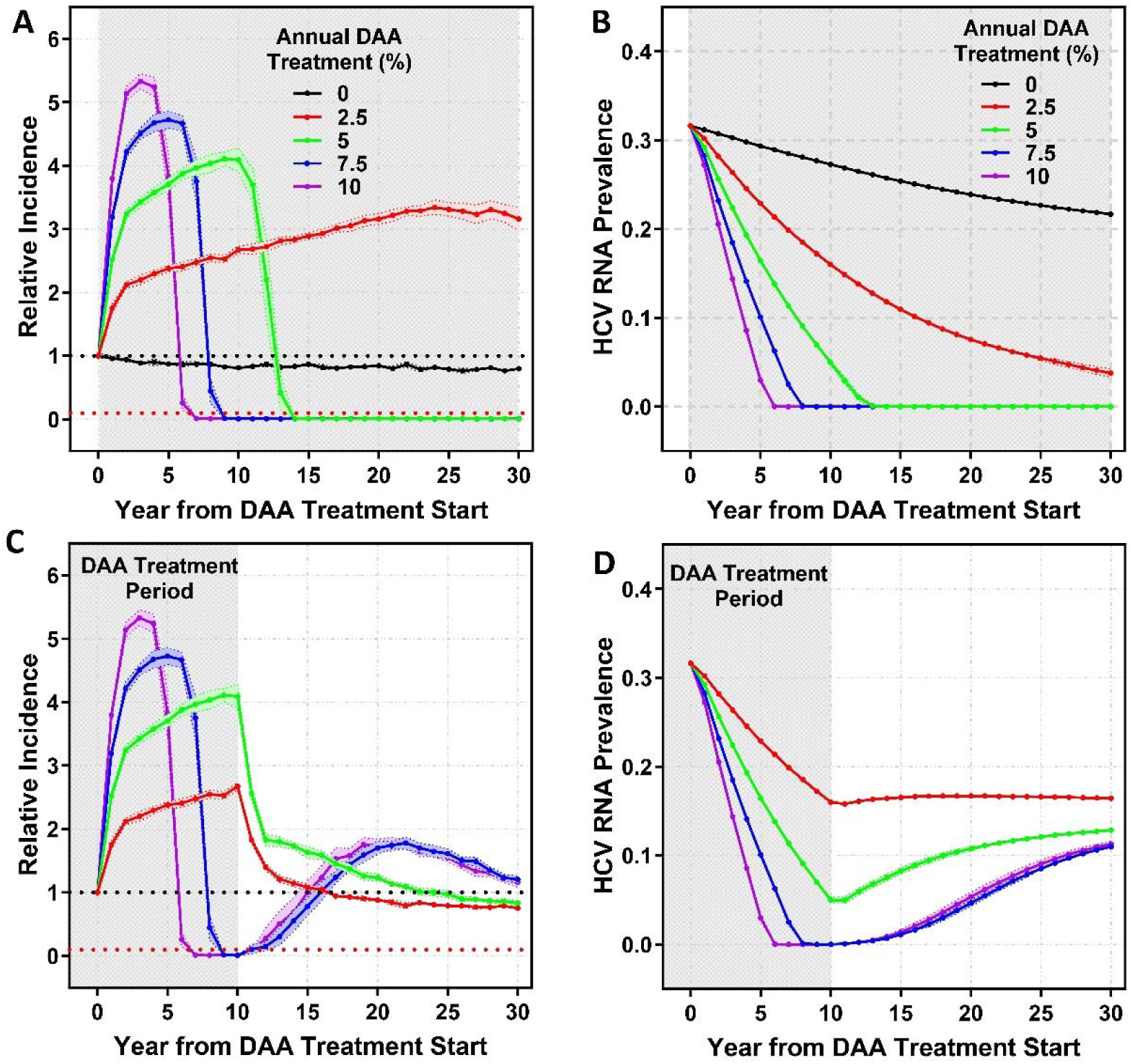
Incidence and prevalence of HCV infections following DAA treatment in the entire PWID population. **(A)** Incidence of new chronic infections **(B)** HCV RNA prevalence. Data are shown relative to the year DAA treatment is initiated (0). **(C)** Incidence of new chronic infections relative to year (0) and after year 10 in which DAA treatment is discontinued. **(D)** HCV RNA prevalence after year 10 in which DAA treatment is discontinued. Line colors in (C) and (D) correspond to initial treatment levels as shown in (A) and (B). Shaded areas represent periods of DAA treatment. Lines and points are the mean values over stochastic replicates and ribbons represent the 95% CI. The black dashed lines represent the pre-treatment incidence (1.0) and the red dashed lines represent the WHO elimination goal (0.1) (**A,C**).

### Target DAA scale up of more than 7.5% is projected to achieve the elimination target within 10 years

For higher than 2.5% DAA treatment rates, we also observed an initial increase in relative incidence (Fig. 3A) before levels began to decline. Modeling indicated that the elimination goal (90% reduction in incidence or 0.1 in Fig 3A) can be achieved within 10 years with a DAA target rate of 7.5% or 10% (Figure 3A). A 7.5% treatment rate achieves the elimination goal within 9 years, and a 10% treatment rate achieves the goal within 7 years. Decreasing the target treatment rate to 5% substantially increases the timeline to elimination to 14 years from the start of DAA treatment (Figure 3A).

Once the elimination goal is met, HCV incidence remains at ∼1% of the pre-treatment level for each of the 5%, 7.5% and 10% groups. HCV RNA prevalence declines for all groups after DAA treatment is started (Figure 3B), with the rate of decline consistent with the treatment rate. Following treatment, prevalence drops to less than 1% within 13 years for rates of 5-10% after 30 years.

### Sustained DAA treatment is required to achieve and maintain HCV micro-elimination among PWID

When DAA treatment is discontinued after 10 years, the incidence of new chronic infections gradually increases for the groups that initially reached the elimination goal (7.5% and 10% treatment) and returns to the pre-treatment baseline within 5-6 years (Figure 3C). Treatment rates of 2.5% and 5% did not achieve the elimination goal within 10 years, but rather increased incidence by 2-4-fold (Fig. 3A and shaded area in Fig. 3C). When treatment is stopped after 10 years, (i) the incidence of new chronic infections gradually declines due to the decrease in new, susceptible PWID that have been treated and returns to the pre-treatment baseline within 25 years (Figure 3C) and (ii) HCV prevalence increases steadily to between 10-20%, but does not reach pretreatment levels (Figure 3D).

### Reduced DAA treatment target is needed to sustain the elimination goal after it is achieved

Rather than stopping treatment completely, we examined the impact of a range of reduced treatment rates for the 7.5% initial treatment group over a 10-year period (Fig. 4). We found that a target annual DAA treatment rate as low as 0.5% (160 individuals per year) is sufficient to sustain elimination (Fig. 4A), maintaining the incidence at ∼1% of the baseline level. Target treatment rates below 0.5% resulted in either rapid increases in incidence to baseline levels within 5-6 years (0-0.1% target treatment rate) or a gradual increase over the subsequent 20 years to ∼60% of the initial incidence (Fig. 4A). Increasing the target DAA treatment rate above 0.5% (1-7.5%) did not result in substantially lower incidence (Fig. 4B) because the number of available infected people to treat remains below the target rate.

**Figure 4.**
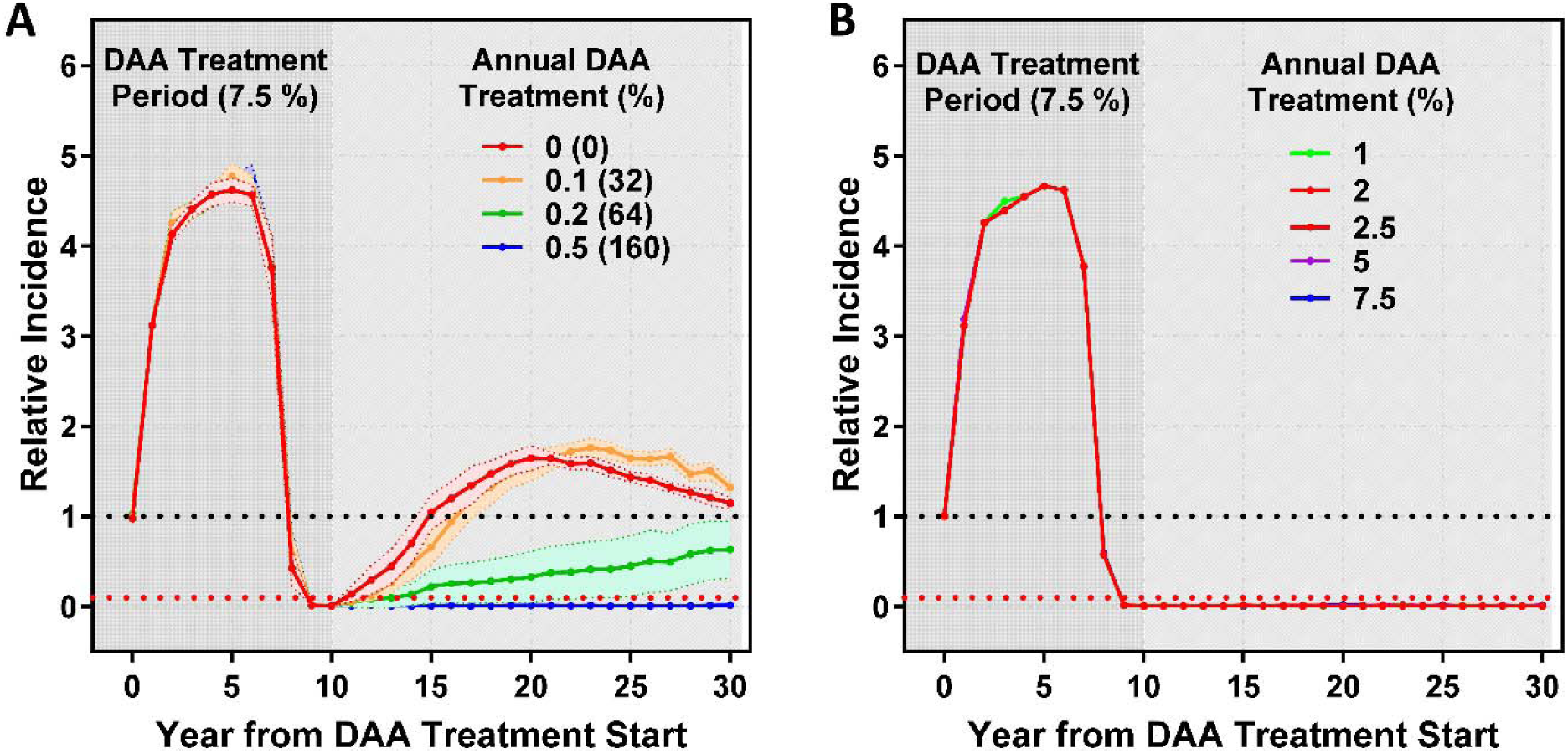
Incidence of new chronic HCV infections when DAA treatment is reduced post-elimination. **(A)** Applying target treatment rates of 0 to 0.5% of the entire population **(B)** Applying target treatment rates of 1 to 7.5%. Data shows incidence relative to the year (0) in which DAA treatment is initiated, and after year 10 in which DAA treatment is reduced for the remaining 20-year simulation timeframe. Dark shaded area represents initial DAA treatment level of 7.5%, light shaded area represents reduced DAA treatment levels. Lines and points are the mean values over stochastic replicates and ribbons represent the 95% CI. The black dashed line represents the pre-treatment incidence (1.0) and the red dashed line represents the WHO elimination goal (0.1).

## DISCUSSION

Direct-acting antivirals (DAAs) against HCV have changed the landscape for treatment of chronically infected people. They are highly effective in achieving virologic cure with 8-12 weeks of treatment [5]. The WHO elimination goal of reducing HCV incidence by 90% has proved difficult in many countries, including the United States, due to limitations in funding and the availability of coordinated treatment programs [7]. We examined several factors associated with HCV elimination using an ABM refined and extended from our previous studies [18, 19] that represents the Chicago, IL PWID population and encompasses drug use and syringe sharing behaviors, demographic and behavioral characteristics, and geographic locations. This complex ABM uses empirical data to form an in-silico population that includes syringe-sharing networks and relationships.

While in our previous HepCEP model we assumed a fixed probability (1%) of transmission through syringe sharing among infected PWID [18], herein we extended HepCEP by incorporating in-host viral kinetics at all stages of infection and treatment (Fig. 2) to allow for a more realistic, dynamic HCV transmission rate using mathematical models of in-host viral kinetics (Supplementary File) to determine transmission probabilities relative to the HCV RNA titers of syringe-sharing donors [22].

Using the extended HepCEP model that accounts for a viral-load-dependent transmission probability, in addition to drug use and syringe-sharing behaviors, networks and geography, we found that for all DAA treatment rates (2.5% to 10%) there was an initial increase in HCV incidence compared to pretreatment levels (Fig. 3A), with the level and kinetics of the increase dependent on treatment rate. This increase was higher than we previously reported [18], in which we assumed a fixed probability (1%) of transmission through syringe sharing, indicating that the use of in-host kinetics impacts predicted transmission outcomes in this population. The observed increase in incidence is assumed to be due to an increase in the numbers of susceptible individuals being generated for each treatment group as PWID are cured of their HCV infection. Chronically infected patients cured of HCV do not have protective immunity, making them susceptible to reinfection with a high probability of developing new chronic infections [13]. Consistent with this hypothesis is the observation that the greatest increase in incidence was seen for the 10% annual DAA treatment group, which would generate more newly susceptible PWID, and the lowest was seen for the 2.5% annual treatment group, generating fewer newly susceptible PWID (Fig. 3A). After the initial increase in HCV incidence, we observed a sharp decline in new cases for the 5-10% treatment groups who went on to reach the elimination goal. In contrast, the HCV incidence for the 2.5% treatment group continued to increase reaching levels 3-fold over the baseline by the 30-year point (Fig. 3A). These findings emphasize the importance of targeting the appropriate number of PWID in a population not only to reach the elimination goal, but also to avoid increasing incidence, which would only become apparent after several years.

Despite the increased incidence observed in the 5-10% treatment groups during the first 3-10 years of treatment and the continued increase in incidence for the 2.5% treatment group (Fig. 3A), we observed a decrease in HCV prevalence for all groups from the start of treatment (Fig. 3B) due to the reduced number of infected people overall in the population. However, the decline in the number of infected people was not sufficient to translate to a reduction in the number of new infections (incidence). This comparison demonstrates that a sharp decline in prevalence is needed to decrease incidence and that only assessing HCV prevalence in a population can be misleading with respect to the rate of new infections.

A critical question regarding elimination of HCV using DAAs is how to maintain the low incidence and prevalence levels achieved after many years of intense and costly treatment. Our modeling found that with complete cessation of DAA treatment upon reaching the elimination goal, HCV incidence returned to baseline levels within 5 years (Fig. 3C). This was associated with an almost 2-fold increase in incidence above the baseline levels over the first 10 years after cessation of DAA treatment, presumably due to the large numbers of susceptible PWID who continued to share syringes. The overlapping 95% CI on incidence following treatment cessation for both the 7.5% and 10% DAA treatment levels (Fig. 3C) indicate that the time to return to pre-treatment baseline incidence is not dependent on the DAA treatment rate. This is expected as the number of infected PWID remaining in the population is the same for both groups regardless of the time taken to reach the elimination goal.

Furthermore, we show that only 0.5% of the PWID population needs to be treated annually to maintain the HCV elimination target once it is met (Fig. 4A). However, this equates to 160 persons that would need to be identified and treated annually within our in-silico population of 32,000 PWID for Chicago, IL and the large surrounding suburban area. Finding this small number of people in a PWID population would be extremely resource intensive. The use of advanced tools, such as theoretical, individual-based models, could contribute to new approaches that would help to overcome the logistical challenges involved in such a major undertaking.

Overall, our findings demonstrate the difficulties in reaching and maintaining the HCV WHO elimination goal without careful consideration of challenges in recruiting and treating PWID, as well as risk practice dynamics within the population. Our findings emphasize the dangers of underestimating annual treatment rates and the potential impact of reduced funding once the elimination goal is met. A previous study used a compartmental model [35] to understand post-elimination dynamics of HCV among PWID. While the model predicted overall similar concerns, the more complex HepCEP model considers individual HCV viral load trajectories and transmission probabilities, in addition to the syringe sharing networks along which the transmissions can occur and has been specifically designed to inform public health intervention goals to reach elimination among PWID. These modeling studies emphasize that continued investment will be critical after reaching the elimination goal. Harm reduction intervention strategies such as development of an HCV vaccine and syringe service programs could complement the public health benefits of DAA treatment in the long term.

Limitations of this study include unbiased random sampling for DAA treatment, which assumes perfect knowledge of HCV infections in all PWID. In practice, identifying infected individuals is costly and time-consuming, particularly when prevalence is low. This could have an impact on the time to reduce incidence and may require higher treatment rates to compensate. Second, a DAA treatment adherence of 90% was used. This may be an optimistic assumption, however our previous modeling studies [18] have shown nominal differences between HCV incidence during DAA treatment as long as there was no limitation placed on the number of DAA treatments each PWID could receive, as we assumed here. We have previously shown that retreatment is essential in PWID populations to achieve elimination goals and that without retreatment the goals cannot be met in a realistic timeframe [18]. Third, our model assumes no limitation on local DAA treatment resources, which could vary widely among different countries and regions. We also acknowledge that this ABM uses a PWID population in metropolitan Chicago. Populations in other areas of the US or the world may have varying access to treatment and harm reduction services, challenges in recruitment, syringe-sharing rates and HCV incidence that require modified treatment rates to achieve the desired goals. The incidence in this in-silico population is lower relative to other metropolitan areas in the U.S., suggesting that the treatment rates would need to be higher in populations with a higher HCV incidence and the impact of underestimating treatment rates could have a greater impact on HCV incidence in the long term. Similarly, the DAA treatment rates needed to maintain reduced incidence could be different depending on the syringe-sharing behaviors of the specific PWID population. This further serves to highlight the complexities in effectively treating PWID populations and underscores the need for other intervention strategies to maintain or achieve the elimination goal, such as syringe services programs and vaccines.

## Data Availability

All data and code used for running experiments, model fitting, and plotting are available on our GitHub repository.

https://github.com/hepcep/hepcep_model

