## Supplementary material for "Modeling highlights the challenge of maintaining HCV micro-elimination among people who inject drugs": Suppl File

**Table of contents**

HCV Infection Dynamics using In-Host Viral Kinetic Models………………… 2-8

Table S1………………………………….…………………………………………………… 5

Model Population, environment and network characteristics………………..9-10

Table S2………………………………….…………………………………………………… 11

Table S3………………………………….…………………………………………………… 12

References…………………………….…………………………………………………… 13-14

***HCV Infection Dynamics using In-Host Viral Kinetic Models***

HepCEP includes a dynamic HCV state transition model for each individual PWID agent that determines HCV infection status after an infection occurs. Previously, HepCEP used parameter estimates for HCV transmission probability and duration of acute and chronic HCV infections state changes, self-clearance, and treatment periods [1, 2]. In the extended HepCEP, we implemented a new state transition using a mathematical model [3, 4] of individual viral load profiles and transmission probabilities based on empirical studies to better reflect the temporal dynamics of viral load and HCV transmission probability unique to each PWID.

When a PWID agent becomes infected, the infection state is categorized as naive infected (NI), meaning not previously infected or cured by DAA treatment, or reinfected (RI) meaning that the agent had cleared infection spontaneously. For each HCV infection profile type, NI or RI, individuals were assigned a viral load profile belonging to one of three groupings for each type (Fig 2, main text) based on our previous work [4].

For acute infection in naive PWID, three viral load profiles were assumed (Figure 1, main text): (*NI1*) full control (spontaneous clearance of HCV RNA within 6 months post infection), (*NI2*) incomplete control (≥1 log10 IU/mL decline in HCV RNA following the peak titer followed by persistence), or (*NI3*) persistence (increase or < 1 log10 IU/mL decline in HCV RNA following the peak titer). In PWID with HCV reinfection (ie., after spontaneous viral clearance) we classified viral load profiles as (*RI1*) rapid clearance (within 4 months based on [4]) after low peak viral titer, (*RI2*) rapid clearance (within 4 months) after high viral titer, or (*RI3*) chronic infection (Fig. 1, main text). To generate synthetic viral load profiles with first-time acute infection or acute infection after developing immune response (re-infection), we performed mathematical modeling of acute infection [3] as described below.

***Viral Load to Probability of Transmission***

We previously developed an approach to translate the likelihood of an infection taking place based on a needle sharing event [4]. This probability relates the amount of virus left behind by the first individual in a needle sharing event, which depends on the viral load of the first individual, the type of needle used and if the needle was rinsed in between uses. Here we assumed that low dead space syringes (LDSS) were used and that these syringes were not rinsed. We then created a probability of transmission function based on the individual’s viral load and used this function to map from an agent’s viral load to the per event transmission probability.

***Mathematical Modelling of In-Host Viral Kinetics***

**Acute Infection**

For the acute infection cases, we obtained the mean and standard deviation of the viral load in infected PWID falling into the categories of clearance, incomplete control, or persistent infection from [4]. We then adapted our model of viral kinetics (VK) [3] to obtain parameter estimate ranges for the given cases. The adapted VK model consists of the following equations:

$$\frac{d}{dt}\left( T \right)=s+rT\left( 1-\frac{I+T}{T_{max}} \right)-dT-\beta VT+q(t)I$$

$\frac{d}{dt}\left( I \right)=\beta VT+rI\left( 1-\frac{I+T}{T_{max}} \right)-{(\delta}_{0}+\kappa(t)+q\left( t \right))I$	(Eq. 1)

$$\frac{d}{dt}\left( V \right)=\left( 1-\varepsilon(t) \right)pI-cV$$

$$q\left( t \right)=\{0, t<t_{kq} q_{0}, t>t_{kq}$$

$$\kappa\left( t \right)=\{0, t<t_{kq} \kappa_{0}, t>t_{kq}$$

$\varepsilon\left( t \right)=\{0, t<t_{\varepsilon} \varepsilon_{0}, t>t_{\varepsilon}$.

*Where T* is the concentration of target cells, *I* is the concentration of productively infected cells, and *V* is the viral load. The VK model assumes that target cells are produced at constant rate *s* and die at constant rate *d*. De novo infection is at constant rate *β*. Infected cells die both from natural attrition and from additional viral-mediated effects at rate *δ_0_*. Furthermore, there is a time-dependent component of the infected cells death rate, *κ(t),* from immune-mediated killing. There is also noncytolytic clearance of virus from infected cells that takes place at rate *q(t)*. Before time, *t_kq_*, *κ = q = 0*, whereas after this time the values increase. Target cells and infected cells proliferate at a maximum proliferation rate *r*, under a blind homeostasis process, with no distinction between infected and noninfected cells. Production of hepatitis C virions by infected cells occurs at average rate *p* per cell and clearance of virions is at rate *c*. There is also a time-dependent component ϵ(t), which controls the production of virions set to 0 before the milestone time *t_ϵ_* and increases to 0 < ϵ < 1 after *t_ϵ_*. The following parameter values were fixed: *T_max_*=1.87×10^7^, *r*=0.01, *d*=0.002, *𝜅_0_*=0.01, *𝜀_0_*=0.94, *𝛿_0_*=0.0036, *s=37,400, c=10*, and *t*_𝜀_=10 days. We also define the parameter V_0_ which was used as a fitting parameter that controls the final viral load of the patient. This was used to set p=1.01E-5*V_0_, 𝛽=0.0936/V_0_. The initial values were set such that at the start time, T(0)= *T_max_*, I(0)=1, V(0)=0.01.

Then the parameters V_0_, *q* and *t_kq_* were fit. The fits were carried out on the mean viral load over time and for one standard deviation above and one standard deviation below the mean. For Incomplete control, the profile was only fit up to the first 6 months of data in order to capture the subsequent rebound as part of the general rebound pattern. Across all 3 profiles of full control, incomplete control, and persistence, these gave overall ranges for V_0_ of [3.99-9.38], for *q* of [0.016-0.379], and for *t_kq_* of [50.0-179.89].

To generate the viral load profiles, we randomly generated 50 sets of parameter values for V_0_*, q*, and *t_kq_* for each profile between the ranges of the maximum and minimum value that the parameters had in the fits for that profile. We then ran the VK model with the chosen random parameter values and saved the results as representing individual PWID viral load time series. The range of values for each profile in the fits is shown in **Table S1**.

|  |  | V_0_ [log IU/ml] | *t_kq_* [days] | *q* | *t*_𝜀_ [days] |
| --- | --- | --- | --- | --- | --- |
| **Acute** | Full Control | 3.94-7.94 | 50.0-61.8 | 0.250-0.293 | 10.0 |
|  | Incomplete Control | 4.46-7.15 | 48.49-100.0 | 0.1-0.13 | 10.0* |
|  | Persistence | 4.24-6.61 | 24.46-84.74 | 0.01-0.05 | 10.0* |
| **Reinfection** | Clearance/ low titer | 1.76-3.88 | 23.87-76.39 | 0.166-0.647 | 10.0* |
|  | Clearance/high titer | 4.49-7.68 | 14.53-124.0405 | 0.192-0.999 | 10.0* |
|  | Chronic | 2.60-6.65 | 14.75-69.04 | 0.031-0.096 | 5.68-12.37 |

**Table S1. Parameter values used to create viral load profiles.** V_0_ is related to baseline value of the viral load, *t_kq_* is the time delay before the increase in immune response leading to cell death of infected cells and the start of non-cytolytic clearance, *q* is the rate of cytolytic clearance and *t*_𝜀_ is the time delay before blocking of production starts. *Parameter was fixed for this case.

**Long-Term Viral Load**

For chronically infected individuals and for individuals experiencing rebound, the long-term value of viral load tends to converge to a fixed value within some range. Our recent reports that include more than 250 chronic HCV-infected individuals who were evaluated before DAA therapy was initiated suggest that the long-term viral load is around 6.071±0.65 log titers [5-8].

To ensure that our VK model produces viral load profiles for chronic HCV infected PWID that reach a viral load steady state of around 6.071±0.65, we extended the VK model with the originally selected parameters up to a randomly selected time, *t_inc_*, between 6 months (180 days) and 1 year (360 days). After that, we draw a value for the parameter V_0-new_ which will replace the original V_0_, which we now refer to as V_0-old_. The value for V_0-new_ is drawn randomly in the range between 6-8 logs, which was empirically found to best correspond to the real data. The increase is assumed to take 6 months (180 days) and takes place according to

$$V_{0}\left( t \right)=V_{0-old}{10}^{\left( V_{0-new}-log \left( V_{0-old} \right) \right)*\frac{t-t_{inc}}{180}} (Eq. 2)$$

until 180 days have passed since the increase began.

After that we have the fixed value V_0_= V_0-new_. The increase in viral load based on Eq. (2) is shown in chronic PWID cases (Figure 2, main text), middle and right).

**Reinfection**.

Only individuals who have experienced spontaneous clearance of a naive acute infection (Profile NI1) can be considered ‘reinfected’ if they experienced infection again because persons who achieve resolution of acute infection produce antibodies that then remain even after the infection is cleared. These anti-bodies change the body’s response to infection a second time. Previous work [4], examined reinfected individuals and also classified them into three groups - those that experience low titers with clearance, high titers with clearance, and chronic infection. Of 27reinfection cases studied, 12/27 experienced low titers with clearance, 5/27 experienced high-titers with clearance, and 10/27 reached chronic infection. We follow those same probabilities in assigning individuals to one of the 3 profiles in our simulation.

To generate the profiles for reinfection, a similar procedure was followed as in naïve acute cases. However, for each of the three aforementioned viral load profiles of reinfection the same model, Eq. (1), was used with the same fixed values as stated above but for the reinfection cases the parameters were fit to individual patient profiles that belong to each of the 3 profiles. For the low titer clearance case, this involves fits for 8 PWID; for the high titer clearance this involves fits for 5 PWID and for the chronic case this involves fits for 10 PWID. As before, we take the maximum and minimum values of the parameters and then draw values for the parameters within that range in order to create *in silico* PWID. Due to poor resulting fits for patients falling under the chronic profile, *t*_𝜀_ was set to be a variable parameter rather than being fixed to *t*_𝜀_ =10. Finally, for the chronic case, we used the same procedure (Eq. 2) as outlined above to ensure that the long-term viral load follows what is expected for chronically infected individuals.

**Direct-acting antiviral (DAA) Treatment**

To model the effect of treatment on infected individuals, data from six studies exploring mathematical modeling of early HCV kinetics under DAA in CHCV-infected patients were gathered [9]. Daily viral kinetics were generated using a biphasic model with the following equations

$$V(t)=V_{0}[A e^{-\lambda_{1}\left( t-t0 \right)}+\left( 1-A \right)e^{-\lambda_{2}\left( t-t0 \right)}] t>t0$$

where $\lambda_{1,2}=\frac{1}{2}((c+\delta)\pm[(c-\delta)^{1}+4\left( 1-\varepsilon\right)c\delta]^{\frac{1}{2}})$ and

$$A=(\varepsilon c-\lambda_{2})/(\lambda_{1}-\lambda_{2})$$

Of the 276 patient profiles generated based on the sample of patients, one was randomly selected for the given individual selected to correspond to viral response of the patient undergoing treatment.

Treated individuals who reach sustained virological response (SVR) are assumed susceptible to reinfection as DAA therapy does not lead to protective immunity [10]. For the case of unsuccessful treatment (i.e., non-SVR), we also incorporate in the ABM the possibility of relapse. To model relapse after their viral load dropped to 0, we assume that the patient remains with undetectable levels of virus for an additional 7 days. After that, we assume an exponential increase of viral load from 0 to 6 logs over 7 days (not shown).

***Model Population, environment and network characteristics***

A. **Hepatitis C Elimination in PWID (HepCEP) model synthetic population**

The PWID population of metropolitan Chicago is modeled based on empirical data [11]. Sources include the Community Outreach Intervention Projects (COIP) syringe service program, the National HIV Behavioral Surveillance study (Chicago site), and other local studies. Attributes of the synthetic population (termed CNEP+, for Enhanced COIP Needle Exchange Program), are shown in **Table S2.** In brief, parameter estimates were generated to profile each of the estimated 32,000 PWID [12] residing in metropolitan Chicago represented in the synthetic population. The select attributes of CNEP+ have been previously published in [2] and are presented again in **Table S2 and S3** for completeness.

B. **Geographic environment and network formation**

In HepCEP, syringe-sharing is modeled as the mode of HCV transmission and PWID are connected via syringe-sharing networks. Network formation is determined by the probability of two persons encountering each other in their neighborhood of residence or within known drug market areas in Chicago, Illinois [13]. The methods used to calculate network encounter rates, establishment processes, and removal of networks have been previously published [2]. The annual turnover rate of the population is about 2%.

**C. HepCEP Model validation**

HepCEP was previously validated and results showed high concordance, i.e., the predicted and actual values match within 2% overall for HCV prevalence [2]. Similarly, data from a 2012-13 network and geographic study [15] of 164 PWID ages 18-30 and their drug-using network members, was used to calibrate and validate the network formation process. The simulated and actual networks matched closely with an average error of only 1.3% [2].

| *Demographic attributes* |  |
| --- | --- |
| Residence | Chicago: 46%; Suburbs: 54% |
| Race/ethnicity | Non-Hispanic (NH) white: 58%, Hispanic: 18%; NH-black: 21%; NH-other: 3% |
| Gender | Female: 30%; Male: 70% |
| Age | Mean: 35.3 years; IQR: 26.1-43.0; Over 30: 59%; Under 30: 41% |
| HCV infection state | Infected (acute or chronic): 30%  Recovered (antibody +): 13% |
| *Behavioral attributes* |  |
| Duration of injection drug use | Mean: 11.4 years; IQR: 3.3-16.0 |
| Probability of receptive sharing Ranges from 0 (never) to 1 (every injection) | Mean: 19%, IQR: 0%-37% |
| Daily drug injections | Mean: 2.5; IQR: 0.89–3.26 |
| Enrollment in any HR program | SSP: 48%; non-SSP: 52% |
| *Network attributes* |  |
| In Degree (receptive network size) | 56% - 0 (no network), 32% - 1, 12% - ≥2 |
| Out Degree (giving network size) | 65% - 0 (no network), 25% - 1, 10% - ≥2 |

**Table S*2*. Attributes of the CNEP+ synthetic population.**

**CNEP+:** Enhanced Community Outreach Intervention Projects (COIP) Needle Exchange Program (CNEP) population generated for HepCEP (Further details can be found in [2]).

NH = Non-Hispanic; IQR = interquartile range; SSP = syringe service program

| Parameter description | Value | Range | Source/Notes |
| --- | --- | --- | --- |
| Probability of chronic infection | 0.67 | 0.49-0.74 | [16, 17] |
| Attrition rate (per year) | 0.024 | 0.01-0.08 | [18] |
| Burn in days | 365 | - | Calibrated by observing the time necessary for the HCV incidence to stabilize. |
| Initial PWID population | 32,000 | 30,000-34,000 | [12] |
| Mean injection career duration (years) | 30.3 | 10-35 | [18] |
| Probability of cessation | 0.232 | 0.13-0.33 | [18] |
| Probability of acute HCV-infected PWID at time of inclusion | 0.05 | 0.01-0.09 | 0.9% prevalence among newer PWID, and 9% among other* PWID [19] |

**Table S3. Parameters for the generation of the CNEP+ synthetic population.**

*PWID who acquired HCV prior to initiating into injection drug use through other modes (e.g. sharing non-injection drug paraphernalia such as snorting straws.

**CNEP+:** Enhanced Community Outreach Intervention Projects (COIP) Needle Exchange Program (CNEP) population generated for HepCEP (Further details can be found in [2]).
